# Polygenic Risk Links to the Gut Microbiome: The MAGI Catalog

**DOI:** 10.64898/2026.05.29.26354433

**Authors:** Oliver Aasmets, Jelisaveta Džigurski, Nele Taba, Merli Koitmäe, Estonian Biobank Research Team, Triin Laisk, Elin Org, Reedik Mägi, Kristi Läll

## Abstract

Polygenic risk scores (PRSs) can effectively identify individuals at risk across various health conditions, yet their association with the gut microbiome remains uncharacterized. We systematically analyzed associations between 4,794 PRSs covering 615 traits and diseases and the gut microbiome within the Estonian Microbiome Cohort (N > 2,500). Microbiome diversity was associated with 62 distinct PRSs across 10 traits, indicating that genetic predisposition is linked to significant alterations in the microbiome composition. At the species level, 282 associations were identified across 100 PRSs for 34 traits, with triglyceride measurements, glucose regulation, and chronotype measurement PRSs showing the strongest signals. Mediation analysis suggests that the microbiome is altered by physiological changes linked to genetic risk but can also mediate this risk. These results help define early microbial biomarkers and explain inter-individual variability. The findings are accessible through the interactive Microbiome And Genomic Interaction (MAGI) Catalog to support future research.

## Main text

The human gut microbiome, encompassing hundreds of microbial species, is a complex ecosystem that can profoundly influence host physiology^1,2^. Its modifiable nature and functional capacity make the microbiome a promising target for precision medicine, allowing, for example, improvements in disease risk prediction^3,4^ and in personalizing medication use^5^ and diet^6^. At the same time, the microbiome composition itself is influenced by various factors, including physical activity, diet, drug use, and disease prevalence, resulting in significant inter-individual variability in its composition^7–9^. In addition, part of this variability is explained by the human genetic variation^10,11^. While individual genetic variants have been reproducibly linked to specific microbial taxa, the large inter-individual variability and limited sample sizes remain limitations for fully capturing the effects of host genetics on microbiome composition^10,12,13^. Polygenic risk scores (PRSs) can address these limitations by aggregating the effects of genetic variants to quantify cumulative genetic liability^14^. Here, we present the first systematic analysis of 4,794 PRSs from the Polygenic Score Catalog^15^ in relation to the gut microbiome (**Figure 1a**). We identify microbiome characteristics associated with genetic risk across diverse disease modalities and investigate the causal architecture of the observed associations. In addition to showing a novel source of variability, these results guide the identification of early microbial biomarkers to improve personalized risk estimation. To facilitate reuse of these results, we developed the Microbiome And Genomic Interaction (MAGI) Catalog, an interactive resource (available at https://magi.gi.ut.ee/) that enables users to explore PRS–microbiome associations across traits, microbial features, and analysis settings. The catalog supports trait-level and species-level queries, visualization and download of association results, and filtering by PRS category, microbial feature type, and sex. By making the full association landscape accessible, MAGI provides a resource for hypothesis generation, replication, and follow-up studies.

**Figure 1.**
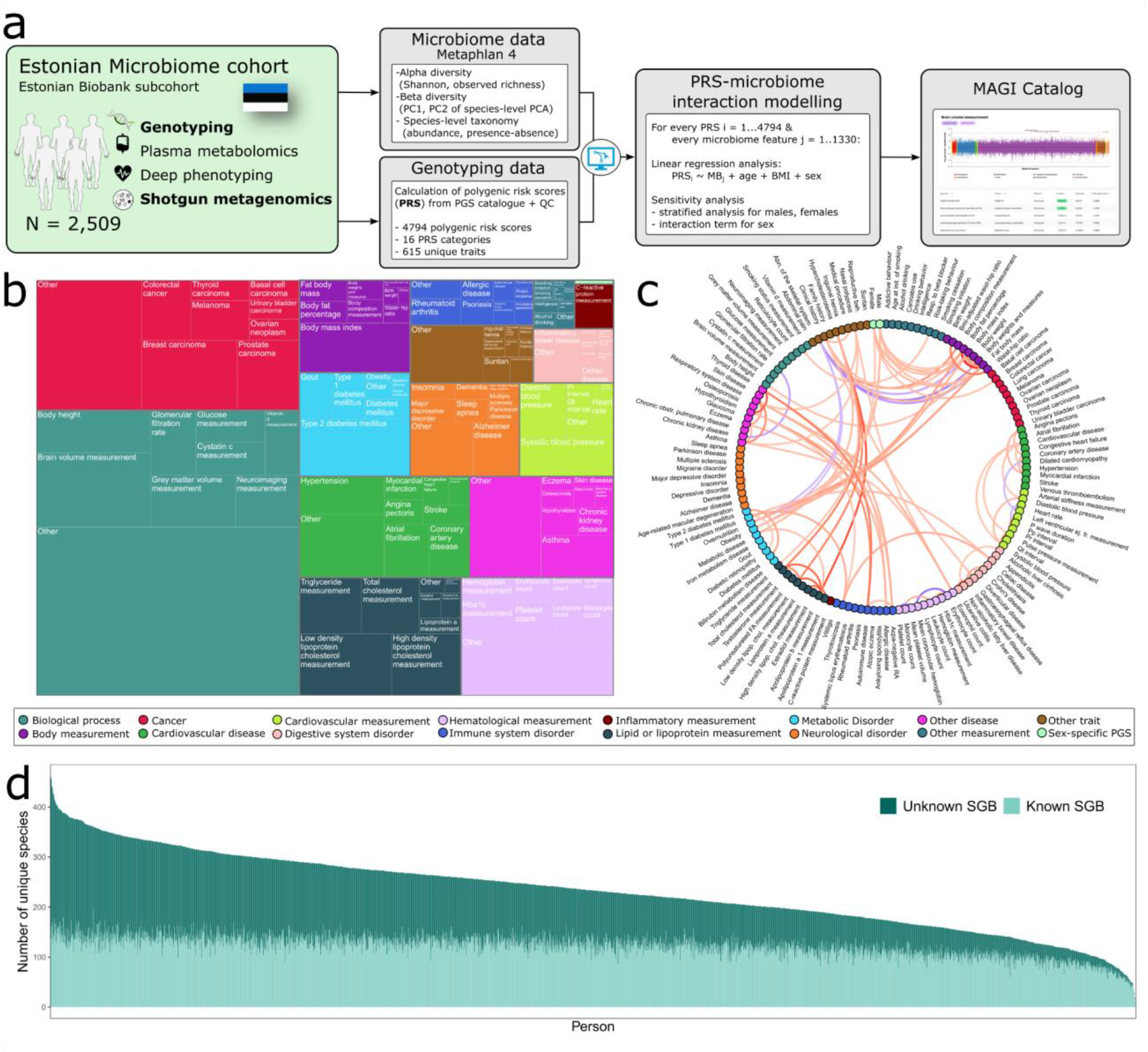
Study workflow & data overview. **a** - Analysis workflow. **b** - Distribution of the 4,794 polygenic risk scores (PRSs) colored by the PRS category. Top 10 mapped traits by the number of unique PRSs per trait are shown for each PRS category. Remaining mapped traits are shown as “Other”. **c** - Average correlations between the PRSs, only shown for abs(cor_avg) > 0.2. The top 10 mapped traits per category according to the number of unique PRSs are shown for each category. **d** - Number of observed microbial species genome bins (SGBs) for the Estonian Microbiome Cohort participants, colored by the reference genome availability for the SGB.

First, we calculated 5,036 PRSs from the Polygenic Score Catalog for 2,509 participants from the Estonian Microbiome Cohort (EstMB)^7^. After removing scores that included Estonian Biobank samples in the development stage or in the underlying genome-wide association study (GWAS)(**Online Methods**), our analysis included 4,794 PRSs. The included PRSs from 16 trait categories (**Figure 1b**) span across 615 unique mapped traits, with the majority of the traits having multiple available PRSs (68.6%; n_max_ = 169 PRSs for brain volume measurement). This reflects the varying scientific and public health interests across traits, differences in endpoint definition, underlying GWASs, and the scoring algorithm used. Moreover, as the PRSs within a trait may highlight distinct pathophysiological mechanisms, we did not further narrow our analysis to a single PRS per trait. The correlations between PRSs for different mapped traits were generally weak and more evident within the same trait category (**Figure 1c**). Microbiome profiling was done using MetaPhlAn4^16^, which defines the species-level genome bins (SGBs) representing both known and unknown species (**Figure 1d**). Analyzed microbial features included alpha-diversity (observed richness and Shannon index), the first two principal components of the SGB-level microbiome profile (PCA on Aitchison distance^17^), and 1,326 SGBs with ≥1% prevalence in EstMB. In the primary analysis, we used the log2-transformed abundance (relative abundance multiplied by the estimated number of cell counts^18^) of the SGBs (**Figure 1a**). Using linear regression, we modeled each PRS against microbial features, adjusting for age, sex, and body mass index (BMI), and deposited the results in the MAGI Catalog.

We identified 62 unique PRSs across 10 traits significantly associated with microbiome diversity (FDR≤0.05)(**Figure 2, Supplementary Table 1)**. Primarily, those associations were present with observed richness (54 PRSs for 6 traits) and the first principal component (50 PRSs for 7 traits), indicating that genetic predisposition to certain traits is associated with broad differences in microbiome composition. PRS traits significantly associated with observed richness included neutrophil and leukocyte counts, chronic lymphocytic leukemia, and triglyceride measurement, all of which were negatively associated (**Figure 2a**). Analysis at the SGB level identified 282 associations for 100 PRSs across 34 mapped traits (FDR ≤ 0.05). The results include 123 unique SGBs predominantly from the *Firmicutes* phylum, mostly associated with one (98 SGBs) or two (24 SGBs) traits. In contrast, the number of significant SGBs per PRS trait ranged from 1 to 40, highlighting that genetic background effects on the microbiome are not only taxa-specific, as also indicated by a recent large-scale microbiome GWAS^12^. PRS traits with the highest number of associations include triglyceride measurements, traits associated with glucose regulation (e.g., glucose measurements, diabetes mellitus), chronotype measurement, skin disease, and thyroid disease (**Figure 2b, Supplementary Table 1**). Notably, when focusing on the triglyceride measurement, the results are consistent with the observation of reduced richness with higher triglyceride PRSs (**Figure 2c**). Analysis identified 10 SGBs whose abundance is reduced with high triglyceride PRS values, thereby allowing specification of the association observed for richness. All 10 SGBs are unknown, suggesting that the effect of genetic predisposition on the gut microbiome may be understudied, and that the development of computational tools and databases is likely to provide further insights.

**Figure 2.**
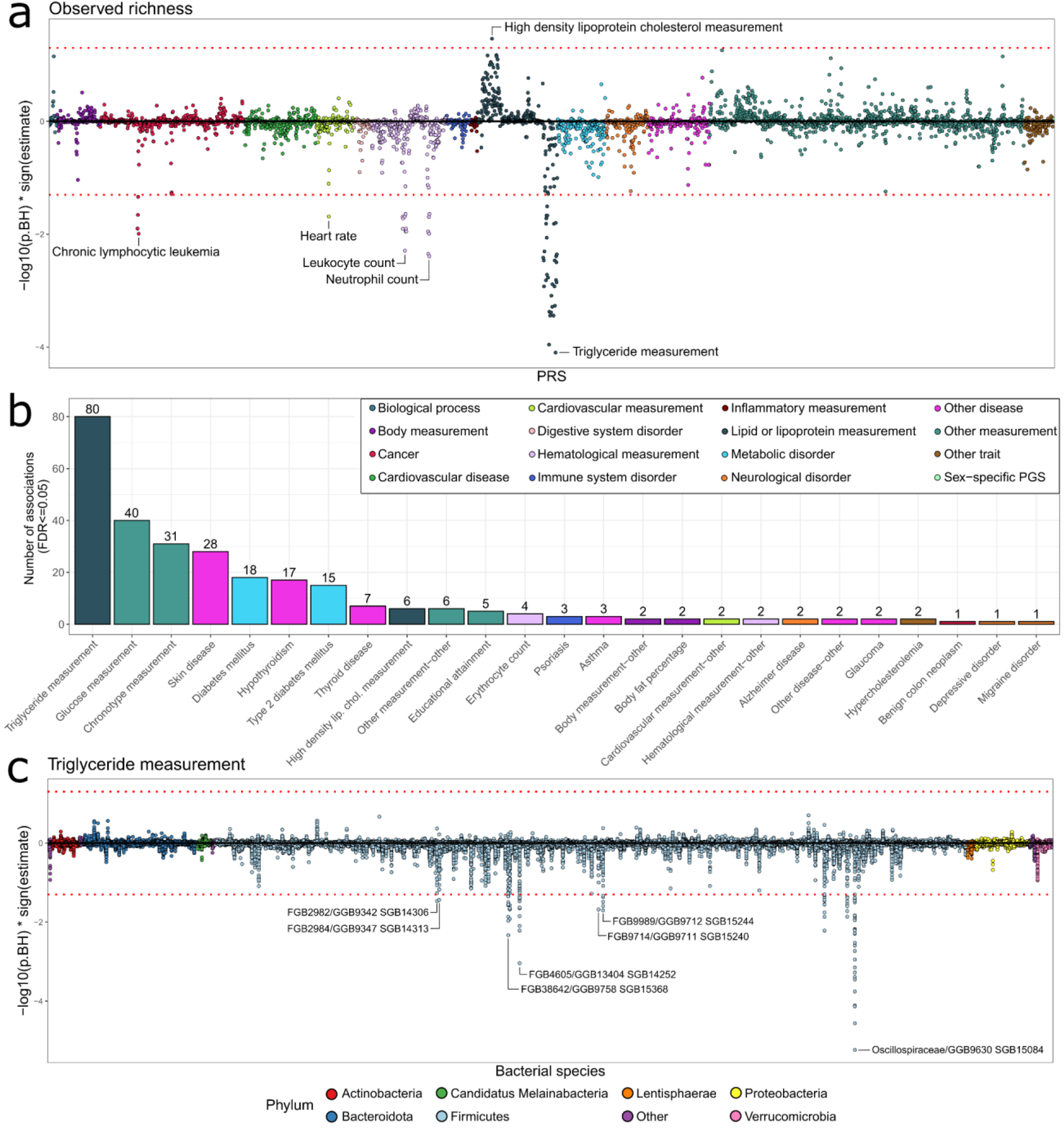
Overview of the main findings. **a** - Associations between PRSs (x-axis) and observed microbial richness. The most significant association per PRS trait is annotated. **b** - Number of FDR hits per PRS trait, colored by the PRS category. **c** - Associations between microbial SGBs (x-axis) and PRSs for triglyceride measurement. The most significant SGB per family is annotated.

We conducted additional sensitivity and subgroup analyses to confirm the robustness of our results. First, analysis of SGB presence-absence (PA) showed consistent signals with the abundance-based analysis. Results of the primary analysis were replicated (n_hits_=171, FDR≤0.05) or at least nominally significant in the PA analysis and with consistent effect directions (**Supplementary Table 2**). This indicates that the identified associations are robust to biases induced by data compositionality. Next, we analyzed interactions with biological sex as complex traits can have a sex-specific genetic architecture^19^. We identified significant microbiome-sex interactions for 9 traits, of which the majority were with PRSs for type 2 diabetes, psoriasis, and intraocular pressure measurement (**Supplementary Table 3**). However, we likely lack sufficient statistical power to fully characterize these interactions. For instance, one PRS for glucose measurement (PGS002924) displays visibly different association patterns in the sex-stratified analysis (**Supplementary Figure 1**; n_females_=1764, n_males_=745), although these differences should be interpreted cautiously given the limited sample size, particularly in males. These observations are broadly consistent with prior reports of sex-specific genetic and PRS associations for type 2 diabetes^19,20^.

The observed associations raise the question of whether, given a genetic background, microbiome composition is altered due to changes in the trait of interest, or whether the microbiome itself is a precursor or a mediator. As a case example, we performed a bidirectional analysis of triglyceride measurements, microbial richness, and *Oscillospiraceae* abundance to understand the causal architecture. First, we conducted a formal mediation analysis, in which the primary model (PRS->Triglycerides->Microbiome) tested whether circulating lipids mediate the genetic effect on the microbiota, while the reverse model (PRS->Microbiome->Triglycerides) tested whether the microbiome mediates the genetic effect on circulating lipids. The primary model emerged as the dominant pathway for both richness and *Oscillospiraceae* abundance, characterized by a substantially higher proportion mediated (Richness prop_primary_=47.6%, Prop_reverse_=4.4%; *Oscillospiraceae* prop_primary_=84.0%, Prop_reverse_=2.5%) and a statistically significant negative average causal mediation effect (ACME; richness mean_ACME_=-3.21; *Oscillospiraceae* mean_ACME_=-0.041; p≤0.05), indicating that genetically driven hypertriglyceridemia likely suppresses microbial richness (**Supplementary Table 4**). This host-to-microbiome effect remained robust in participants not taking statins, confirming an intrinsic biological response rather than a medication artifact. A significant average direct effect (ADE; Richness mean_ADE_=-3.63; p≤0.05) for richness in the primary analysis suggests that the genetic architecture of lipid metabolism also influences microbiome richness through pleiotropic pathways independent of circulating triglyceride levels. This was not consistently observed for *Oscillospiraceae*. However, a statistically significant ACME was also observed in the reverse model for both richness and *Oscillospiraceae* abundance, suggesting a reciprocal feedback loop where the microbiome further reinforces lipid dysregulation (**Supplementary Table 4**). Interestingly, *Oscillibacter*, which is a member of the *Oscillospiraceae* family, has previously been causally linked to decreased triglyceride concentrations^21^. This is in line with our findings and indicates that a genetic predisposition to higher triglyceride levels can suppress Oscillospiraceae abundance, which, in turn, might further increase triglyceride levels. Taken together, our findings suggest that host genetic liability to elevated triglycerides primarily influences microbiome composition, while allowing for secondary microbiome-mediated effects, highlighting a complex, potentially bidirectional host– microbiome interaction. We also assessed the potential causal effect of triglycerides on microbiome richness using a two-sample Mendelian Randomization approach. However, the results remained inconclusive, possibly due to insufficient statistical power and a small sample size in the GWAS of microbiome richness. Therefore, although follow-up studies are needed to validate these findings, our results can guide the generation of causal hypotheses and support inference.

Taken together, the observed significant correlations with genetic risk and partial mediation through the microbiome imply that alterations in the microbiome can precede future health deterioration. Thus, our results may highlight conditions in which the microbiome can serve as an early biomarker. A similar approach based on *GBA1* gene variants recently identified microbial biomarkers for the development of Parkinson’s disease^22^. We believe that the broad scope of available PRSs can significantly advance the detection of such microbial biomarkers. These results also show that information about disease risks can be partially shared between genomic and microbiome data. Thus, the development of risk scores combining these data layers should take it into account. Therefore, although further analysis is required to systematically map the underlying causal pathways, our results can inform future studies and provide clinical translation.

Despite these advances, several limitations should be considered. First, although this represents, to our knowledge, the first systematic analysis of gut microbiome associations with a wide spectrum of genetic predispositions, the available sample size limits the detection of smaller effect sizes. The relatively high proportion of nominally significant associations suggests that additional PRS–microbiome relationships likely remain undetected. For example, PRSs for urolithiasis, household income, and keratoconus showed an excess of nominal associations, suggesting that genetic liability for these traits may also influence microbiome composition. Increasing sample size will be essential to improve statistical power, refine effect estimates, and more clearly resolve both overall and sex-specific associations. Second, replication remains a key challenge in this field. Although numerous large-scale biobanks and clinical cohorts exist, many have already contributed to GWASs and the development of PRSs. As the same cohorts begin to generate microbiome data, sample overlap may limit the availability of fully independent datasets, complicating external validation and potentially inflating observed effects^23^. Missing information about cohorts included in studies in PGS or GWAS catalogs can also contribute to the given issue and lead to artificially high associations due to undetected sample overlap. Double-checking top associations for PRSs could mitigate the risks. However, it is not feasible to manually double-check all PRSs or GWASs. In addition, the transferability of externally derived PRSs can constrain the detection of robust signals, particularly across populations with differing genetic architectures. Consistent with this, several PRSs with significant associations in our study showed reduced single-nucleotide variant overlap (<75%) with the original score profile, potentially limiting their portability and replication in other cohorts. Finally, because microbiome profiles were measured cross-sectionally and are influenced by medication use, diet, lifestyle and disease status, the observed associations should primarily be interpreted as links between genetic liability and microbiome variation rather than direct causal effects.

In conclusion, our findings imply that analyzing host genetic information summarized as PRSs can improve our understanding of the effects of genetic predisposition of a trait or disease on the gut microbiome. The results, made available through the MAGI Catalog, provide input for future studies and enhance method development and causal inference in microbiome research.

## Online Methods

### Study population

The Estonian Biobank (EstBB) is a volunteer-based biobank established in 2001 that today includes approximately 212,000 participants of predominantly European ancestry, representing about 20% of Estonia’s adult population^24^. Participants signed broad informed consent, and for these individuals, biospecimens and extensive health, demographic, and biological data have been collected via questionnaires and regular linkage to various Estonian national health databases.

The Estonian Microbiome cohort (EstMB) was established in 2017 when stool, oral, and blood samples were collected for a subset of 2,509 EstBB participants^7^. All participants in the EstMB cohort provided informed consent for their data and samples to be used for scientific purposes. A detailed overview of the EstMB, including omics and phenotypic data availability, has been described in Aasmets & Krigul *et al*. 2022^7^.

### Genotyping and imputation

Estonian Biobank participants were genotyped using four versions of the Illumina Global Screening Array (GSAMD-24v1, GSAMD-24v2, ESTchip-1_GSAv2, and ESTchip-2_GSAv3) across 17 genotyping batches, each comprising at least 1,000 samples. Quality control (QC) was performed for each batch separately. At the sample level, individuals with a call rate below 95% or a mismatch between genetically inferred and reported sex were excluded. At the variant level, genotypes were removed if they failed any of the following criteria: call rate <95%, Hardy–Weinberg equilibrium exact test p-value <1×10^−4^, Illumina cluster separation <0.4, or GenTrain score <0.6. All genotyping data were aligned to the GRCh37 human genome reference. To account for potential batch effects, variants with inconsistent allele frequencies across batches were excluded. Allele frequencies were calculated for each batch containing more than 10,000 samples (n = 9), and a mean frequency across these batches was derived. Variants showing a deviation greater than 5% from the mean in any batch were removed.

Only single-nucleotide variants (SNVs) were retained for imputation. Ambiguous strand variants (A/T and G/C) and rare variants (minor allele frequency <1%) were excluded. For SNVs genotyped by multiple probes, concordance between probes was evaluated within each sample, and discordant genotype calls were removed on a per-sample basis. A second round of variant-level filtering (call rate ≥95%) was then applied. Following QC, all batches were merged into a single dataset. In total, approximately 307,000 SNVs passed all QC filters and were used for downstream phasing and imputation. Phasing was performed without a reference panel using Eagle (v2.4.1). Imputation of the phased data was carried out using Beagle (v5.4; *beagle*.*22Jul22*.*46e*.*jar*) with a local copy of the Haplotype Reference Consortium panel, comprising 27,165 reference samples and including only polymorphic sites with at least five minor alleles.

### Polygenic risk score calculation

For all EstMB participants, polygenic risk scores (PRSs) were calculated using the PGS Calculator (pgsc_calc v2.0.0-alpha.2) tool and scoring files for all traits and diseases available in the PGS Catalog up to March 2025^15^. No imputation quality or MAF thresholds were applied to variants before calculations, and the calculated PRSs were not adjusted for individuals’ genetic ancestry, as participants were mainly of European ancestry^24^. Of the 5,036 scoring files downloaded, only 15 could not be calculated due to formatting incompatibilities with the pgsc_calc tool or to large variant missingness in the scoring file (overlap<0.001). In the following steps, all PGSs (n=242) that included EstBB samples in the development stage or in the underlying genome-wide association study (GWAS) were removed, resulting in 4,794 PRSs available for downstream statistical analyses. According to the accompanied PRSs metadata, such as reported and mapped trait name, all PRSs were categorized into 16 categories: biological process, body measurement, cancer, cardiovascular disease, cardiovascular measurement, digestive system disorder, hematological measurement, immune system disorder, inflammatory measurement, lipid or lipoprotein measurement, metabolic disorder, neurological disorder, other disease, other measurement, other trait, and sex-specific PGS. For each score, we extracted from the *report*.*html* file provided by *pgsc_calc* the column “Match %” and determined the overlap between the original score SNV list and SNVs available in EstBB data.

### Metabolomics

The metabolomic data were generated using nuclear magnetic resonance (NMR) technology from Nightingale Health, and NMR measurements were conducted on EDTA plasma samples (100 μL) for all EstBB participants. Data collection, quality control, and preprocessing have been previously described in detail^25,26^. Here, we used the log-transformed total triglyceride measurements in the mediation analysis.

### Metagenomic data creation

#### Microbiome sample collection and DNA extraction

The participants collected a fresh stool sample immediately after defecation with a sterile Pasteur pipette and placed it inside a polypropylene conical 15 ml tube. The participants delivered the sample to the study centre, where it was stored at −80°C until DNA extraction. Microbial DNA extraction was performed using the QIAamp DNA Stool Mini Kit (Qiagen, Germany). Around 200 mg of stool was used as starting material, following the manufacturer’s instructions for the DNA extraction kit. DNA was quantified from all samples using a Qubit 2.0 Fluorometer with dsDNA Assay Kit (Thermo Fisher Scientific). The NEBNext® Ultra™ DNA Library Prep Kit for Illumina (NEB, USA) was used to generate sequencing libraries according to the manufacturer’s recommendations. Briefly, 1 μg DNA per sample was used as input material. Index codes were added to attribute sequences for each sample. The DNA sample was fragmented by sonication to an average size of 350 bp. DNA fragments were end-polished, A-tailed, and ligated with the full-length adaptor for Illumina sequencing with further PCR amplification. Finally, PCR products were purified (AMPure XP system), and libraries were analyzed for size distribution using Agilent2100 Bioanalyzer and quantified using real-time PCR. **Metagenomics annotation**. Shotgun metagenomic paired-end sequencing was performed by Novogene Bioinformatics Technology Co., Ltd. using the Illumina NovaSeq6000 platform, resulting in 4.62 ± 0.44 Gb of data per sample (insert size 350 bp, read length 2 x 250 bp). The reads were trimmed for quality and adapter sequences. The host reads that aligned to the human genome were removed using *SOAP2*.*21* (parameters: -s 135 -l 30 -v 7 -m 200 -x 400)^27^. Taxonomic profiling was performed using MetaPhlAn4^16^ with default parameters. As a result, relative abundance was obtained for 2,692 species-level genome bins (SGBs).

### Statistical analysis

#### Microbiome data preprocessing

To assess alpha diversity in the gut microbiome, observed richness and the Shannon diversity index were used. The Shannon index was calculated using the *vegan* package v2.5-6^28^. To assess microbiome beta-diversity, principal component analysis (PCA) was performed, and the first two principal components were used for the analysis. Euclidean distance on the centered log-ratio (CLR) transformed microbiome SGB-level profile was used to calculate between-sample distances for the PCA^17^. To apply the CLR transformation, zero counts were imputed with a pseudocount equal to half of the minimal non-zero relative abundance value. For SGB-level analysis, we included SGBs present in more than 1% of the EstMB samples, yielding 1326 SGBs. For the primary analysis, the relative abundance was multiplied by the estimated cell count^18^ and log2-transformed. As a sensitivity analysis, the presence/absence of the SGB was assessed, with 0 used as the threshold for SGB presence.

#### Primary association analysis

To assess associations between each microbial species and each PRS, linear regression models were used, adjusted for sex, age at microbiome sampling, and body mass index (BMI). Specifically, in a manner analogous to a phenome-wide association study, we performed a systematic pairwise analysis in which, for each PRS (i = 1,…,4794) and each microbial feature (j = 1,…,1330), the following model was fitted:

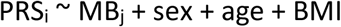

where MB_j_ represents the abundance of microbial feature *j*. To account for multiple testing, we adjusted the p-values using the Benjamini-Hochberg procedure. Analysis was also carried out separately for males and females, and the results were made available in the MAGI Catalog.

#### Sensitivity analysis

To evaluate potential sex-specific effects, sensitivity analyses were conducted by incorporating interaction terms between sex and microbial feature across all PRS–species pairs as follows:

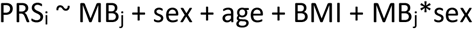

#### Mediation analysis

To assess the causal directionality between the gut microbiome and triglyceride levels, we performed bidirectional mediation analysis using the *mediation* R package (v4.5.0). We used a triglyceride-specific PRS as the exogenous exposure across two frameworks: a primary model (PRS −> Triglycerides −> Microbiome) and a reverse model (PRS −> Microbiome −> Triglycerides). Both the mediator and outcome models were constructed using linear regression, adjusting for age, sex, and statin usage to control for confounding. The Average Causal Mediation Effect (ACME), Average Direct Effect (ADE), and Proportion Mediated were estimated using quasi-Bayesian Monte Carlo simulations with 1,000 bootstrap iterations. To ensure robustness of the host-to-microbe direction, a sensitivity analysis was conducted by restricting the dataset to participants who did not receive statin therapy. All continuous variables were standardized prior to analysis. We ran the analysis separately for each PRS significantly associated with the microbiome trait and aggregated the statistics across PRSs.

#### Mendelian Randomization

To disentangle the detected association between triglycerides-PRS and microbiome richness and attempt to evaluate the directionality of the results, we assessed the potential causality of triglycerides on microbiome richness using two-sample Mendelian Randomization (MR) analysis implemented with *TwoSampleMR* R package (v4.5.1)^29^. The genetic instruments were defined based on GWAS of triglyceride measurements in the UKBB (N = 441,016)^30^. The single-nucleotide polymorphisms (SNVs) with effect-allele frequency <0.01 or >0.99 were removed, and only genome regions of interest were retained prior to performing linkage disequilibrium (LD)-pruning using the *clump_data()* function in the *TwoSampleMR* package. Three different settings were used for designing the instrument-sets: 1) *LPL* gene region with 100kb window on each side, p<1*10^−6^, r^2^<0.1; 2) *LPL, APOC3, ANGPTL3, ANGPTL4* gene regions with 100kb windows on each side, p<1*10^−6^, r^2^<0.1; 3) genome-wide, p<5*10^−8^, r^2^<0.001. The gene regions were defined as GRCh37/hg19 by Ensembl. For the same SNVs, estimates for microbiome richness were obtained from a recent GWAS conducted in a sample comprising four Swedish population-based studies (N = 16,017)^12^. After data harmonization, we had 24 instruments for the *LPL* region, 63 instruments for the *LPL, APOC3, ANGPTL3*, and *ANGPTL4* regions combined, and 278 instruments genome-wide. The inverse-variance-weighted method was used as the main MR method, while MR Egger, MR-RAPS, weighted median, and weighted mode were used for sensitivity analyses. The results were visually assessed using a scatter plot with fitted lines for the causal effect estimates and a funnel plot.

### MAGI Catalog web application

To support exploration of results from conducted analyses and diverse research interests, we developed, as inspired by PheWeb2^31^, the interactive Microbiome And Genomic Interaction (MAGI) Catalog web application. Therein, all analysis results can be queried at the trait- or microbial feature-level, sorted by statistical significance, visualized, and downloaded. The results can also be filtered by PRS trait categories, microbial feature preprocessing (abundance (ABS)/presence-absence (PA)), and analysis subgroup (male, female, combined). Only one PRS (PGS004919) was filtered out because it was flagged as a duplication and retired from the PGS Catalog during application development. It is important to note that, while the PGS Catalog is a continuously updated real-time database, our database is anchored to specific releases, with the first release tied to the information used in this study. Still, all PRS identifiers link directly to the PGS Catalog, where the most up-to-date information about PRS can be found. To improve plot rendering performance, non-significant points were thinned into a single representative point per ~3-pixel grid cell. All statistically significant points, visual density, and peaks were preserved, and unfiltered results are available in the accompanying tables. The MAGI Catalog is available at https://magi.gi.ut.ee/.

## Data availability

The metagenomic data generated in this study have been deposited in the European Genome-Phenome Archive database (https://www.ebi.ac.uk/ega/; data set accession code EGAD00001008448, study accession code EGAS00001005900) and are available upon request. The phenotype data include sensitive information from electronic health registers. Access to pseudomized data from the Estonian Biobank (EstBB) is granted in accordance with the Estonian Human Genes Research Act, and data access occurs via the University of Tartu’s secure analysis environment (SAPU server). Detailed instructions for submitting an application to access Estonian Biobank data are available at https://genomics.ut.ee/en/content/estonian-biobank (see the “Data Access” tab for detailed steps). In brief, the process starts with a preliminary inquiry to, followed by a research application to the Scientific Advisory Committee (SAC) for scientific evaluation.

## Acknowledgements

The authors would like to thank Mari-Liis Tammesoo, Marili Palover, Anu Reigo, Neeme Tõnisson, Liis Leitsalu, Triinu Temberg, and Esta Pintsaar for participating in the sample collection process. We thank Steven Smit, Rita Kreevan, and Martin Tootsi for the DNA extraction process. Data analysis was carried out in part in the High-Performance Computing Center of the University of Tartu.

## Contributions

O.A., M.K., and K.L. designed and T.L., E.O., R.M. and K.L. supervised the study. The EstBB Research Team collected and provided the genotype, metabolome, and phenotype data. O.A., J.D., N.T., M.K., and K.L. performed and interpreted the analyses. J.D. developed the MAGI web application. O.A., J.D., N.T., M.K., and K.L wrote the manuscript. All authors discussed the results and approved the final manuscript.

## Ethics declarations

The activities of the EstBB are regulated by the Human Genes Research Act, adopted in 2000, specifically to regulate the operations of the EstBB. In 2026, a new Human Genes Research Act entered into force, establishing an updated regulatory framework for both the EstBB and genetic data–based research in Estonia more broadly. All participants have signed a broad consent form. The use of EstBB and EstMB data for this study was approved by the Ethics Committee of the University of Tartu (No. 266-T10) and the Estonian Committee on Bioethics and Human Research of the Ministry of Social Affairs (No. 1.1-12/2768), and data access was granted by the EstBB under approval No. 6-7/GI/10486.

## Supplementary Figures

**Supplementary Figure 1.**
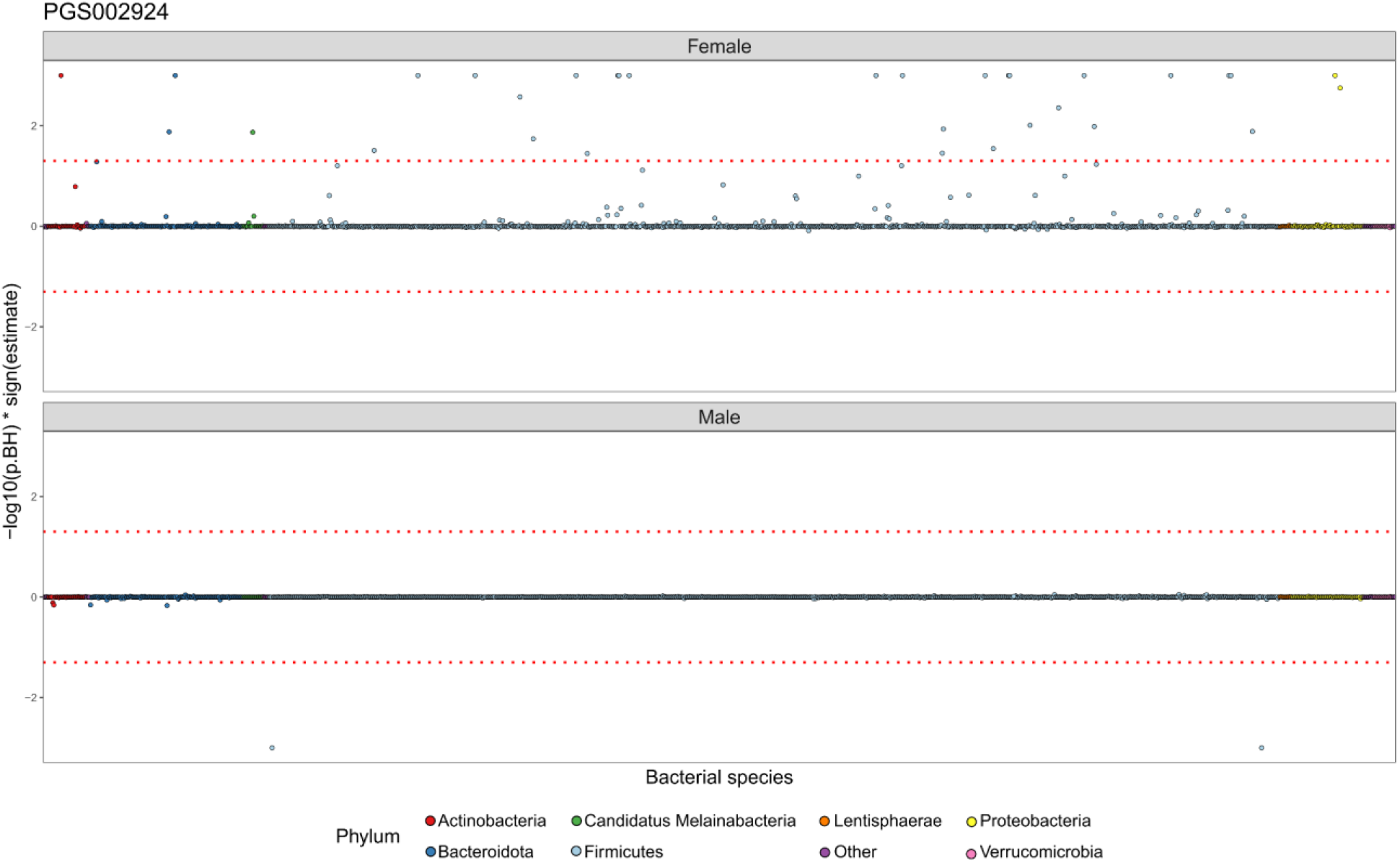
Associations between microbial SGBs (x-axis) and PRS PGS002924 for females and males separately. The y-axis is truncated at −log10(p.BH)*sign(estimate) = +/−3.

